# A Mixed-Methods Evaluation of Clinician Experiences and Adoption Patterns of an EHR-integrated Generative AI-based Clinical Decision Support in Kenya

**DOI:** 10.1101/2025.08.14.25333740

**Authors:** Christopher Obong’o, Grace Njenga, Dickson Otiangala, Edith Jepleting, Sue Wairimu, Mira Emmanuel-Fabula, Sarah Kiptinness, Robert Korom, Brian Taliesin, Bilal A Mateen

**Author notes:** **<u>Corresponding Author:</u>** Dr. Christopher Obong’o, PATH Kenya, ACS Plaza, 4th floor, Lenana Road, PO Box: 76634-00508, Nairobi, Kenya.

## Abstract

**Objective:** To quantify the adoption pattern of an LLM-based clinical decision support system across private primary health facilities in Kenya (operated by Penda Health), and explore factors influencing clinician uptake and overall experience.

**Methods:** A mixed-methods study combining quantitative analysis of CDSS metadata from all consultations that took place between 1^st^ February and 1^st^ October 2024, augmented by qualitative data from 42 staff members (26 clinical officers, 10 facility managers, four nurses, one quality assurance manager, and one business analyst). Data collection included journey mapping interviews (n=7), user-experience interviews (n=25), focus groups (n=2), and system utilization metrics. Quantitative data were summarized using descriptive statistics, and qualitative data analysed using thematic analysis (drawing on established theories of technology adoption and change management).

**Results:** In total, there were 258,106 clinical episodes across all Penda Health facilities over the 8-month observation period, of which 56,050 (21.7%) were augmented by use of the ‘AI Consult’. ‘AI Consult’ use, aggregated across the 16 facilities, increased from 4% to 47% over 8 months. Feedback on the CDSS guidance was infrequent (only being provided in 31% of cases), but when it was, it was overwhelmingly positive (99.5%). The qualitative investigation identified five key themes associated with clinicians’ experiences with the AI Consult tool: (1) there are several value propositions to an ‘AI Consult’ style tool, (2) Clinicians’ confidence in the AI consult grew with time, (3) clinicians’ application of the ‘AI consult’ is influenced by case complexity, (4) responses from the AI consult are largely believed and valued by clinicians but several improvements are recommended, and (5) clinicians find the ‘AI consult’ easy to use but identified several pain points that warrant attention.

**Discussion:** Successful GenAI/LLM-enhanced CDSS implementation in resource-constrained settings requires: (1) robust technological infrastructure, (2) localisation to reflect clinical guidelines, (3) structured change management with clinical champions, and (4) seamless workflow integration. Future product development exercises should specifically consider alternatives to active solicitation of CDSS input, as it is liable to overconfidence-related underutilization.

**Key Questions Box (BMJ Digital Health and AI):** *What is already known?:*

- AI-powered clinical decision support systems face implementation challenges, especially in low-resource contexts.
- Studies from high-income countries suggest that workflow integration and perceived usefulness are key drivers of adoption.
- Limited research exists on the implementation of GenAI in resource-constrained healthcare environments.

*What are the new findings?:*

- It is possible to achieve non-trivial adoption of GenAI-based CDSS (4% to 47% over 8 months) when supported by structured change management.
- There are several value propositions for a GenAI-based CDSS, including improved quality of care and serving as a learning aid.
- Several factors influenced utilization/engagement, from perceived case complexity to ease of use and trust in the system.
- Clinicians’ behaviour and interactions with GenAI tools (as demonstrated by varying prompt use) mature over time based on perceived usefulness.

*What do the new findings imply?:*

- Sustainable AI implementation in low-resource settings requires addressing both technical infrastructure and human factors.
- Relying on active solicitation of inputs from an AI system is likely to suffer from overconfidence bias, which limits its overall utility, and instead, we should explore other methods of integrating CDSS into the relevant clinical workflows.

## Introduction

Generative Artificial Intelligence (GenAI) has emerged as a potentially transformative tool in healthcare, offering advancements in diagnostic accuracy, treatment optimization, and patient-centered care (Goh et al., 2024; Tu et al., 2025). While research on GenAI implementations in high-resource settings is rapidly advancing, and an increasing number of effective solutions are being identified (Heinz et al., 2025) the evidence base for the effectiveness of AI generally is significantly sparser where it concerns resource-constrained environments (Han et al., 2024). A consistent theme that has emerged across the research that does exist for (GenAI-enabled) clinical decision support systems (CDSSs) is that technical accuracy is not sufficient to ensure impact (Ciecierski-Holmes et al., 2022). There are also critical ‘human factors’ requirements, spanning: trust, system usability, workflow integration, adequate training, and supportive organizational culture, that determine uptake and impact (Ciecierski-Holmes et al., 2022; Nii et al., n.d.; Obe Destiny Balogun et al., 2023).

Research from high-income settings suggests that trust, a critical human factor requirement for successful implementation of a novel AI-based product, is bolstered when decision support systems are explainable and clearly assist clinicians’ goals, whereas “black-box” models can undermine confidence (Tun et al., 2025). Interestingly, the evidence around other human factors such as the impact on error occurrence, time requirements, and alert sensitivity (often cited as a major issue due to the risk of alarm fatigue (Olakotan & Yusof, 2020) is more heterogeneous (Lambert et al., 2023). However, misfit with clinical workflow integration is consistently a hindrance to success (Wang et al., 2021), whilst training is an effective way to improve acceptance (Trivedi et al., 2009).

In low-resource primary healthcare settings, the same human factors appear to be important, but they are compounded by context-specific challenges (Adepoju et al., 2017; Manyazewal et al., 2021; Peiffer-Smadja et al., 2024) – in no small part because the nature of the workforce is different (Peiffer-Smadja et al., 2024), with much greater reliance on community health workers or ‘clinical officers’ (degree-trained individuals, who are not medical doctors). For example, limited digital literacy and scarce training opportunities mean that without substantial user support, new decision support tools may be perceived as an added burden rather than a benefit (Ciecierski-Holmes et al., 2022). In fact, health workers often feel overwhelmed by AI systems introduced without sufficient orientation, leading to low uptake unless gradual onboarding and “social learning” are facilitated (Hengstler et al., 2016). Moreover, skepticism that undermines uptake arises when AI tools are not attuned to local practice patterns (Ciecierski-Holmes et al., 2022), as AI-CDSS that fail to accommodate local languages, guidelines, and disease profiles can feel irrelevant or untrustworthy to frontline providers. In sum, while factors like healthworker trust and usability are universally important, low-income settings especially require AI-CDSS solutions to be context-adapted and backed by robust training and evidence, so that providers view them as trustworthy aids rather than foreign impositions.

To develop our understanding of how human factors influence GenAI-specific CDSS product uptake in low resource settings, an understudied issue, we carried out a mixed-methods assessment of a Kenyan primary health care social enterprise’s (Penda Health) implementation of an electronic healthcare record system-integrated, large language model (LLM)-based, clinical decision support system (hereafter referred to as the ‘AI Consult’). In this study, we sought to explore:

1. Patterns of adoption of the ‘LLM Consult’ over time, and across different user groups;
2. The archetypal user journey and how the LLM-based CDSS integrated into routine care; and
3. Factors influencing clinicians’ decisions to use or not use the available LLM consult.

## Methods

### Study Setting

This mixed-methods study was conducted at Penda Health facilities, a private outpatient healthcare provider operating 16 facilities in Nairobi and Kiambu County, Kenya, serving all socioeconomic segments of society (incl. many people from informal settlements), and all age groups (from neonates as young as two days old to elderly patients).

Each staff member is equipped with access to Penda Health’s in-house custom-built EMR system (Easy Clinic, Kolkata, India), into which the LLM (GPT4)-based CDSS is integrated, i.e., the ‘AI Consult’. As illustrated in Figure 1, the ‘AI Consult’ is a single-button feature which results in certain documented material from the EMR being passed to the LLM, accompanied by one of four prompts (selected by the clinician). The high-level description of the prompts and their purpose is detailed below:

- Comprehensive Consult: This prompt provides several paragraphs of feedback to the clinician regarding differential diagnosis, diagnostic testing, history and physical examination gaps, and guideline-based treatments. It’s more in-depth and typically includes a "clinical pearl" for professional development. The purpose of this prompt was to provide a more in-depth analysis of the case with nuanced background and clinical teaching. See the supplementary material for the complete prompt.
- Summarised Consult - The summarised consult was created in response to early clinician feedback that the outputs were too long for busy clinical practice. The summarised consult response provided no more than a paragraph of feedback to the clinician, focusing on actionable steps a clinician could take to improve quality of care.
- Management Plan - The Management Plan prompt was meant to allow clinicians to easily access evidence-based treatment guidelines for the diagnosis or diagnoses listed. The prompt would return a management plan for the condition without focusing on other aspects of care, like the comprehensive and summarised consult prompts.
- How’s my documentation? - This prompt was created in order to ensure clinicians improved the quality of documentation for history and physical exam since all other aspects of AI consult are dependent on the quality of this documentation. It would provide a rating and constructive feedback points based on whether the clinical notes documented were adequate for the condition at hand.

**Figure 1:**
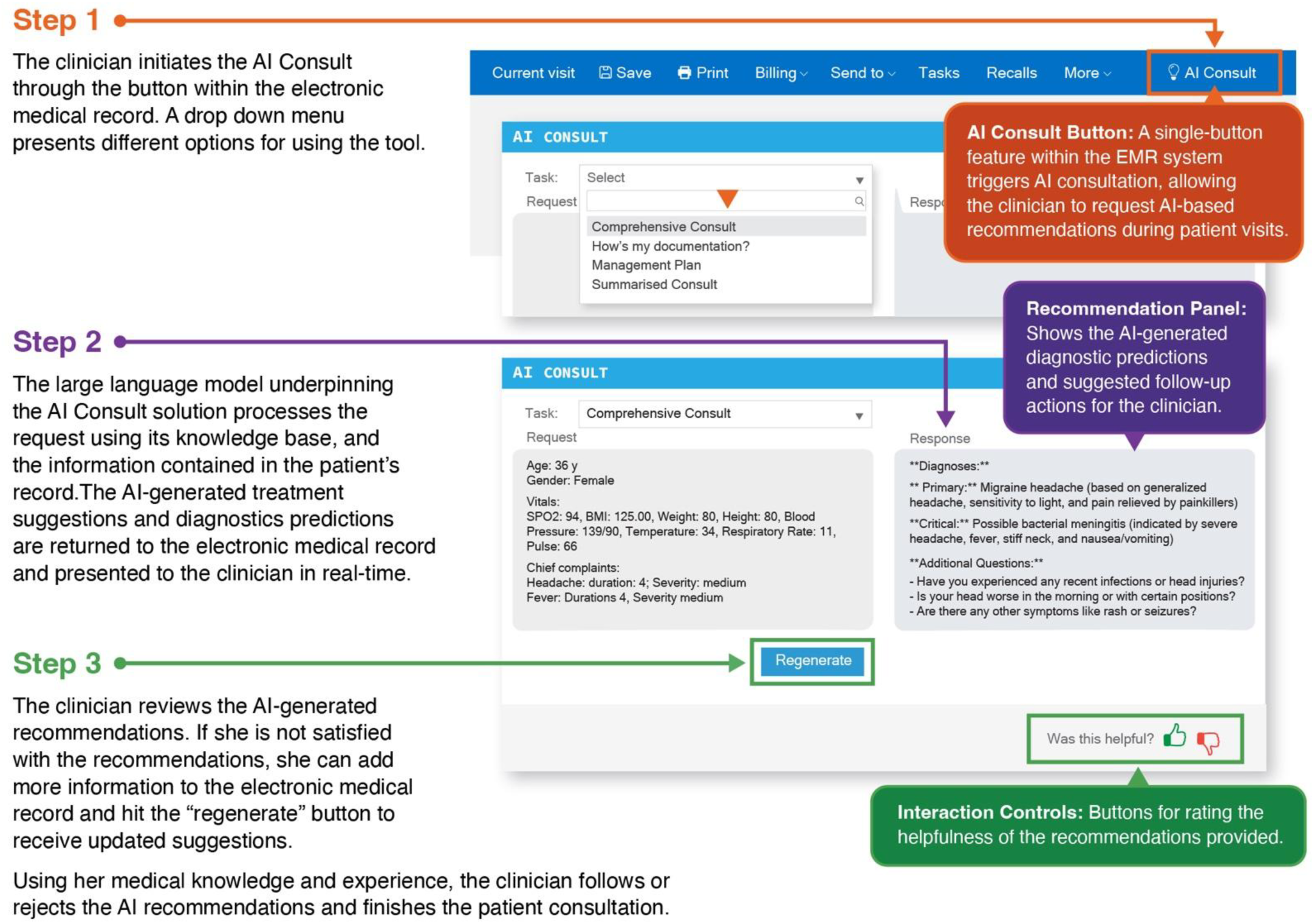
A Summary of the ‘AI Consult’ Workflow.

For all prompts, the totality of de-identified data for the current visit was shared with the LLM. Depending on what stage of the visit the AI consult was used, this could include up to and including:

- Demographic data (e.g., Sex, Age)
- Vital Signs
- Clinical notes (free-text history and physical examination)
- Chronic illnesses
- Pregnancy /Obs/Gyn history if applicable
- Lab tests and results
- Diagnosis
- Treatment (Medication orders)

No fine-tuning of the LLM was undertaken, nor RAG-framework implemented for this version of the ‘AI Consult’.

### Study Design & Data Collection

#### Quantitative Assessment of Uptake

System logs for all consultations completed between 1^st^ February and 1^st^ October 2024 were extracted from the backend database utilised by Penda Health, by their systems administrator, and shared with the research team in an Excel file in December 2024. Each system log contained: (1) a unique ID for the clinical episode, (2) a flag denoting whether the AI consult was utilized, (3) the specific prompt selected by the clinician, (4) medical center where the visit occurred, (5) the visit date (6) the total number of times the AI consult prompts were used during the visit, and (7) the clinician’s feedback (i.e., thumbs up or down, in response to the LLM outputs).

#### Qualitative Assessment of Clinician Experiences/Perceptions of the ‘AI Consult’

The qualitative component of this mixed-methods study involved the application of a set of human-centered design tools and processes (PATH, 2025), to gather clinicians’ experiences and perceptions of the AI consult and to explore factors influencing clinicians’ decisions to use or not use the AI consult. The study team conducted structured observations of clinical processes, one-on-one interviews and focus group discussions with clinicians from a sample of Penda Health facilities.

Eight of the sixteen Penda Health facilities were selected for sampling of staff for the qualitative HCD activities. These facilities included a mix of high and low patient volume units as well as high and low AI consult utilization sites to provide a comprehensive understanding of why adoption varied. Participants (n=42) were randomly selected from these sites (Etikan, 2017), to include clinical officers (n=26), Penda Health facility managers (n=10), nurses (n=4), quality assurance manager (n=1) and business analyst (n=1).

The qualitative data collection methods included:

`o` Journey mapping sessions (n=7 interviews) in three Penda Health facilities examining six key touchpoints in the AI-enabled consultation workflow: login, patient data entry, AI consultation initiation, recommendation review, documentation, and follow-up. Journey mapping sessions were audio-recorded and analyzed using interaction analysis methods (Jordan & Henderson, 1995).
`o` In-depth interviews (n=25) focusing on usability, workflow integration, and perceived impacts. This sample size was determined sufficient for saturation; thematic saturation was reached after 18 interviews and confirmed with 7 additional interviews that yielded no new themes.
`o` One Focus group discussion workshop was conducted with Penda health facility managers (n=10) to validate the findings from the journey mapping and individual interviews.

Interviews and focus groups were audio-recorded, transcribed, and transferred to an analysis miro board in readiness for analysis.

##### Data Management and Analysis

Qualitative data were thematically analyzed using Braun & Clarke’s six-phase framework (Braun & Clarke, n.d.). Two researchers independently coded all transcripts, with themes developed through consensus meetings and validated through member checking with 12 of the interview participants. The number of interviewees who reported a specific belief/theme and the proportion are reported. Quantitative system data were analyzed using SPSS to describe adoption trends over time, and usage patterns.

##### Ethical Approval & Data Governance

This study was conducted under a pre-existing umbrella IRB approval (ref: MSU/DRPI/MUERC/00899/20), for human-centered design research undertaken by PATH’s Living Labs team, from the Western Institutional Review Board (WIRB) and Maseno University’s Ethics Review Committee (MUERC). All participants provided informed consent, and data were anonymized to ensure confidentiality. Raw data and analytical outputs are stored in compliance with local data protection regulation. To address potential power imbalances, interviews were conducted by researchers not affiliated with Penda Health. The study adhered to the STROBE guidelines for reporting observational studies and the SRQR guidelines for reporting qualitative research.

## Results

In total, there were 258,106 clinical episodes across all Penda Health facilities between 1^st^ February and 1^st^ October 2024, of which 56,050 (21.7%) were augmented by use of the ‘AI Consult’ (See Figure 2). ‘AI Consult’ use, aggregated across the 16 facilities, increased from 4% to 47% over 8 months (Figure 2).

**Figure 2:**
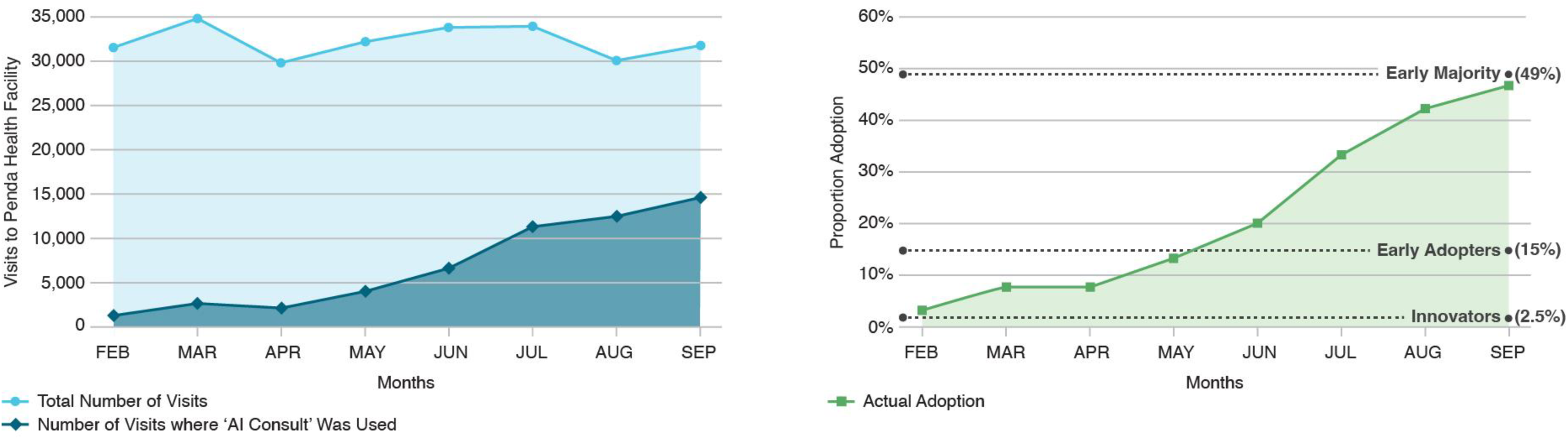
Adoption of the ‘AI Consult’ Across All Penda Health Visits. *(Left) The graph shows (in light blue) the total number of Penda Health facility visits per month, and (in dark blue) the number of visits in which the ‘AI Consult’ was used. (Right) The graphs show the proportion of visits in which the ‘AI Consult’ was used, with superimposed adoption rate thresholds defined by* Rodger’s ‘Innovation-Adoption’ Curve (Everret M, 2014).

Of the 56,050 clinical episodes augmented by use of the ‘AI Consult’, there were 75,519 interactions with the LLM. In 97.6% of cases, these interactions occurred in ‘General Visits’, rather than in ‘Dental’ (35 interactions), ‘Lab’ (220 interaction), ‘Pharmacy’ (5 interactions), ‘Telemedicine’ (921 interactions), ‘Ultrasound’ (75 interactions), or ‘Well Baby’ checks (565 interactions). Moreover, in 43% of cases, the “AI Consult” was used more than once during a clinical episode. The maximum number of times the LLM was used in any clinical episode was 17 times, but it was rare for it to be used more than twice (occurring in less than 17% of cases).

Overall, users strongly preferred the summarized prompt (n = 46,240 of the total 75,519 interactions, 61%) over the management plan (n = 17,631 of the total 75.519 interactions, 23%), the comprehensive consult (n = 11,400 of the total 75,519 interactions, 15%), and the documentation feedback (n = 248 of the total 75,519 interactions, 0.33%). As illustrated in Table 1, by the latter months of the piloting period (February to September), 68% of the (56,050) visits to Penda Health were using the summarized prompt, and the ‘’How’s my documentation" prompt, only introduced in September, was being used minimally.

**Table 1:** Total Number of Monthly Visits Using Each Prompt Type.

| Months | Summarized | Comprehensive | Management Plan | How's My Documentation |
| --- | --- | --- | --- | --- |
| Feb | 1,116 | 305 | 46 | - |
| Mar | 1,379 | 530 | 1,178 | - |
| Apr | 1,094 | 506 | 1,164 | - |
| May | 2,032 | 928 | 1,582 | - |
| Jun | 3,572 | 1,914 | 2,253 | - |
| Jul | 7,664 | 1,991 | 3,165 | - |
| Aug | 9,441 | 1,944 | 2,727 | - |
| Sep | 11,637 | 1,731 | 3,052 | 200 |
| **Totals\*** | **38,384** | **9,904** | **15,299** | **218** |

\*Differences between the sum of the months and the total is due to the study date range being inclusive of October 1st.

Clinicians provided feedback on some interactions; 31% of AI responses had a thumbs-up or thumbs-down rating. Of the interactions with responses, 99.5% were thumbs up, the rest thumbs down.

### Mapping ‘AI Consult’ Use in Routine Care

The high-level (summary) clinical workflow derived from the seven-user journey mapping sessions, and confirmed using the focus group, is illustrated in Figure 3. Within the clinical workflow at the Penda Health facilities, the ‘AI Consult’ is primarily utilized in the consultation room, for challenging or unclear cases, to confirm diagnoses, suggest treatments, or guide treatment plans. The results suggest that there are numerous entry points for the use of the ‘AI Consult’, and as demonstrated later through the qualitative interview analysis, the differences in when the tool was used were often related to features of the clinical case, and the confidence of the clinician in managing it.

**Figure 3:**
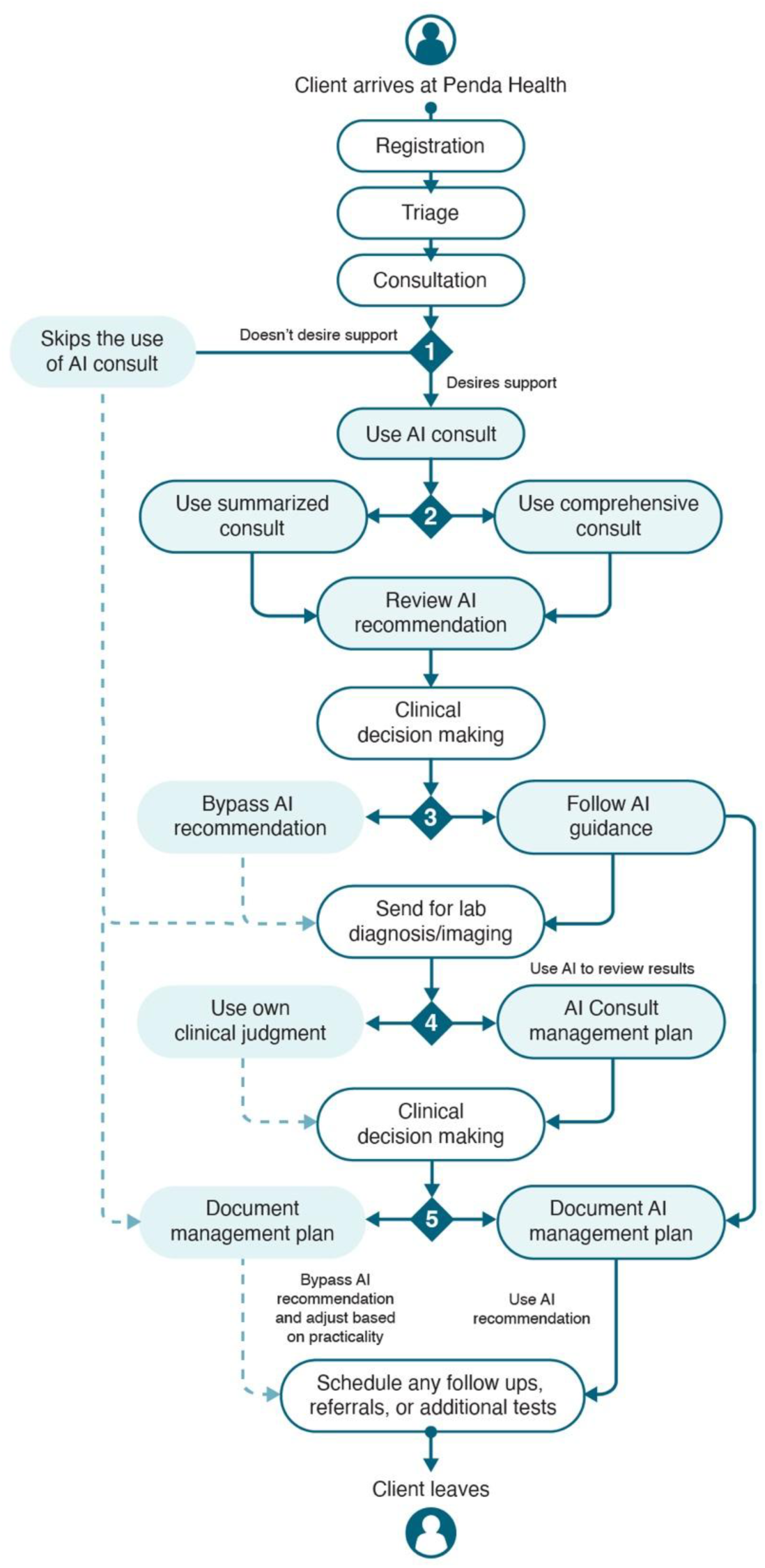
Journey Map of ‘AI Consult’ Use. The first decision is whether to use the AI Consult. If the case appears familiar after gathering initial information, the clinician might choose not to use the tool. If the clinician decides to use the AI Consult, she must then select the appropriate prompt to use. At this stage, most clinicians found themselves choosing between the comprehensive or summarised prompt, and as noted earlier, by the end of the study, the former was almost completely abandoned in favour of the latter. The third decision comes after reviewing the recommendations generated by the AI Consult. Here the clinicians used their knowledge and experience to determine whether to follow the recommendations or bypass them. The fourth decision occurs only if a patient is sent for a test or medical imaging, and in this case, the clinician determines whether to use the tool to review the results. The fifth decision takes place when the AI Consult returns recommendations, and the clinician must then decide whether to use the AI-generated recommendations or create her own treatment/management plan. In some cases, decisions 4 and 5 are bypassed as a clinician feels comfortable after making a management plan after decision 3.

### Clinician’s Experiences and Perceptions of the ‘AI Consult’

Qualitative analysis and synthesis of the interviews identified five key themes:

#### 1) There are several value propositions to an ‘AI Consult ‘style tool

Many of the clinicians reported that the AI consult plays a complimentary role in their clinical decision-making by providing additional insights and constructive feedback on diagnosis, history taking, and treatment plans (n=20).

> *“In most cases, it effectively complements clinical decision-making, provided that a thorough patient history is taken.”* Clinician Penda Health

In other cases, clinicians felt that the consult supports more thorough patient management by prompting additional or overlooked steps, highlighting abnormal vital signs, and suggesting alternative treatments, including non-pharmacological options (n=13).

> *“We mostly use the AI consult in ANC, where clinical details like the chief complaint are documented. Since AI relies on proper input, it becomes especially useful after the provider’s assessment—guiding investigations, management, and even differential diagnoses. It’s particularly helpful during initial ANC visits.”* Nurse Penda Health

Several respondents also noted that the AI consult was useful for learning purposes, providing insights that enhance their clinical skills and understanding (n=8).

> *“I felt it was a valuable addition because it doesn’t limit your thinking or thought process; instead, it offers helpful suggestions. One of the best things about it is that it provides information based on what you’ve input or documented, which broadens your perspective. Overall, it has been a great complement to my clinical reasoning.” Clinician Penda Health*

Notably, one clinician pointed out that there was a noticeable increase in patient satisfaction during the deployment period:

> *"The system has improved patient outcomes significantly. There is a metric called NPS (Net Promoter Score), which gauges how patients feel based on the feedback they provide. The score reflects how patients feel after receiving treatment, and we’ve seen a noticeable improvement in these scores compared to before."* Clinician Penda Health

The net promoter scores for the period in question (and details on how this standardized report data is collected) are included in the supplementary material. The data illustrate an approximate 3.5% increase in NPS from a 3-month rolling average of 73.67 to 77% from the beginning to the end of the observation period (Table s1).

#### 2) Clinicians’ confidence in the AI Consult grew with time, in part due to organisational change management

Clinicians reported experiencing skepticism and fear during the initiation phase of the AI consult (n=7). They feared that the AI consult might replace them as service providers and were skeptical about the added value of the consult in their clinical decision-making.

> *“So while we once thought AI could replace expertise, we’ve come to see that it only enhances your work if you already have the right foundation. The AI makes the process easier, but it doesn’t replace the need for clinical judgment and skills.” Clinician Penda Health*

However, with ongoing training, peer support, and usability improvements over time, perceptions shifted, and trust in the AI consult tool grew; notably, this shift in perception likely underpins the previously described increase in uptake/use.

> *“We had multiple training sessions before the introduction of the AI system. First, the physicians guided us through the training, followed by the IT team. In total, we attended more than three training sessions.”* Clinician Penda Health

#### 3) Use of the ‘AI Consult’ was influenced by case complexity

While some clinicians suggested relying on the ‘AI Consult’ across multiple tasks (n=18), others were more selective (n=6), choosing to rely on the AI consult only for complex or ambiguous cases.

> *“I usually use the summarized consult. Most of the time, I attend to patients using the summarized version because it saves time. However, for learning purposes and complex cases, the comprehensive consult is more effective.”* Clinician Penda Health

A few others indicated they choose to use the ‘AI Consult’ occasionally, often to confirm their decisions (n =10).

> *“I usually use the comprehensive consult to support deeper clinical thinking.” Clinician Penda Health*

In general, application of the ‘AI Consult’ seemed to be more likely for clinicians handling cases perceived to be complex, chronic or needing a second opinion.

#### 4) Responses from the ‘AI Consult’ were largely believed and valued by clinicians, but both hallucinations and a lack of local knowledge (whilst rare) undermined the tool

Clinicians valued the response from the ‘AI Consult’ (n=25), suggesting that it provided accurate guidance most of the time (n=14).

> *“The AI Consult generally provides accurate suggestions. The only issue is that it sometimes recommends medications that aren’t available in Kenya or tests that aren’t commonly done at our facility level*.
>
> Clinician Penda Health

They attributed cases of discrepancies and inaccurate responses from the ‘AI Consult’ to instances where it failed to consider local resistance patterns or context-specific guidelines. For consistent and more confident application of the AI consult, they recommend closer alignment between the AI consult with Kenyan MoH protocols; this would ensure alignment with local practice standards and improve accuracy of responses generated.

> *“AI Consult might recommend actions that slightly differ from the Kenyan protocol, such as fluid management during dehydration or medication use in diabetes, which can be concerning for clinicians familiar with local guidelines.”* Clinician Penda Health

In addition to recommending actions inconsistent with local MoH guidelines, clinicians also reported cases where the ‘AI Consult’ misidentified medications (n=4).

> *"For example, it may classify a cough syrup like Zefcolin [the brand name of a local cough syrup] as an antibiotic, which is incorrect. These errors highlight the need for better recognition of brand names and drug molecules."* Clinician Penda Health

#### 5) Clinicians found the ‘AI Consult’ easy to use but identified several pain points that warrant attention

Some clinicians (n=9) felt the AI consult potentially increases consultation time (for detailed data entry and system processing), while others felt it has reduced the time spent seeking second opinions from colleagues (n=13).

> *"Using AI consult does not increase consultation time, it still remains the same. It actually reduces the time spent to move from room to room to consult a colleagues which used to happen a lot" Clinician Penda Health*

> *“Does it consume time? Yes, to some extent. When you’re not using AI, everything comes straight from your mind, it’s like doing simple math in your head versus using a calculator. For example, one plus one is instantly two when done mentally, but keying it into a calculator takes a bit more time. So yes, using AI may take slightly more time, but not significantly” Clinician Penda Health*

One of the key pain points identified included unreliable internet connectivity that frequently caused delays in receiving ‘AI Consult’ feedback.

> *“It’s easy to use overall. The challenging part is when there’s no network or the system hangs, that’s when it becomes difficult to use.” Clinician Penda Health*

Finally, whilst the clinicians felt that the integration of the AI Consult in the facility’s EMR system enabled the consult to pull data from patient records in providing clinical recommendations and helped improve trust in the responses from the consult, some (n=9) suggested that this integration created an unhelpful dependence on the EMR functionality.

> *“We access it through the EMR, which is where the patient information is drawn from. Whatever is entered into the EMR is integrated with the AI system.” Clinician Penda Health*

## Discussion

This mixed-methods study identified that Penda Health reached 50% utilisation of their AI Consult tool at 8 months post-introduction (i.e., 50% of all visits had at least one recorded use of the AI Consult), with a significant preference for a single prompt style, and specific points along the patient journey when the tool tended to be used. Notably, a non-trivial proportion of physicians chose to circumvent the use of the tool due to their own perceived confidence in managing the clinical complaint they were presented with. The qualitative investigation identified five key themes associated with clinicians’ experiences with the AI Consult tool: (1) there are several value propositions to an ‘AI Consult’ style tool, (2) Clinicians’ confidence in the AI consult grew with time, (3) clinicians’ application of the ‘AI consult’ is influenced by case complexity, (4) responses from the AI consult are largely believed and valued by clinicians but several improvements are recommended, and (5) clinicians find the ‘AI consult’ easy to use but identified several pain points that warrant attention.

### Results In the Context of the Literature

Our findings align with previous studies that highlight the value proposition of AI tools for clinical decision making. Clinicians’ perceptions of the ‘AI consult’ value in improving diagnosis, history taking and treatment plans provide nuanced insights into how an ‘AI consult’ could contribute to “advanced patient care” and “risk reduced patient care”, or “treatment optimization and personalized medicine” and “patient monitoring” in health care settings as documented in the systematic reviews by (Hennrich et al., 2024; Khalifa et al., 2024) respectively.

Previous studies have shown the significance of clinicians’ perceptions of ease of use in the adoption of AI-based clinical support systems (Gonzalez-Smith et al., 2022). Our research builds on these findings by highlighting the value of training and ongoing peer support in shifting initial scepticism and fears to unlock clinicians’ beliefs and adoption of the system. Further, Horwood et al., 2023 reported similar resistance patterns among healthcare workers in South Africa, while Manyazewal et al., 2021 and Oladipo et al., 2024 similarly identified perceived usefulness as critical for AI acceptance in resource-constrained settings.

Finally, consistent with our findings, other studies have documented the educational and learning benefits of CDSS (Khalifa et al., 2024; Singh et al., 2024). However, concerns have been raised about over-reliance on the ‘AI consult’ due to a learned dependance on technology (Goddard et al., 2014; Sutton et al., 2020). We did not explore dependance in this study, but it is an important risk to note that appears under-studied based on the paucity of literature describing the impacts of withdrawal of the technology on clinician quality of care, patient outcomes, or even proximal measures such as clinician job satisfaction and efficiency.

### Strengths & Limitations

Several imitations of this study are worth noting. First, we relied on a relatively modest sample of 42 clinicians drawn from only half the participating facilities. However, the reported results are those we extracted, having achieved saturation in individual interviews, and confirmed through the focus groups, suggesting that they are representative of the population in question. That said, as a for-profit social impact primary healthcare provider, Penda Health’s facilities are not representative of most facilities in Kenya – they have much more robust infrastructure and digital maturity. It is entirely possible that key barriers were overlooked as a consequence, but even in this context, technical infrastructure issues emerged as barriers, suggesting that although barriers are somewhat mitigated at Penda’s facilities, they are not entirely resolved and thus still reflect the broader Kenyan primary care experience.

There are also several strengths to this study; the use of independent interviewers and a separate research team means that we are less likely to have been influenced by the preconceptions of the product development and implementation teams. Moreover, the combination of both qualitative and quantitative data helped triangulate specific behaviours and insights, such as the preference to particular prompts as well as the potential risk of over-confidence biases undermining tools that require clinicians to actively solicit inputs (demonstrated by the non-trivial non-engagement even at 8 months).

### Implications for Product Developers and Policy Makers

Drawing on the results of the analysis, below we present a series of recommendations for implementing the AI consult in similar settings. These insights should inform future product development exercises and efforts to integrate and scale GenAI-based technology in primary health care settings in low- and middle-income countries.

1. Technical infrastructure assessment should precede implementation, ensuring reliable electricity infrastructure, sufficient physical assets (e.g., laptops, tablets, etc), robust internet connectivity, and contingency plans for technical disruptions.
2. Clinical care must be substantially digitized, including biodata, vital signs, clinical history and physical examination, diagnostic test ordering and results, and medication order entry for the data set to be rich enough for a GenAI-based tool to act on.
3. Localization of clinical guidelines is essential for clinician trust and effective use. Implementing organizations must strike a balance between recommending medical best practices and local resource availability.
4. Workflow integration should minimize additional steps for clinicians, recognizing also that active solicitation of support from a CDSS is likely to suffer from overconfidence biases wherein clinicians don’t request support even though it might have been helpful. Version 2.0 of the AI Consult (described elsewhere in Menon et al., n.d. explicitly incorporates this insight.
5. Training and support should be ongoing rather than one-time events. Continuous learning opportunities, responsive technical support, and regular system improvements based on user feedback are critical for sustained adoption.

In essence, healthcare organisations implementing similar systems should consider conducting readiness assessments addressing both technical infrastructure and organisational change capacity before implementation. Phased implementation with regular evaluation can help address emerging challenges and optimize adoption.

## Conclusion

By conducting this mixed-methods UX study in a resource-poor setting, this study extends existing knowledge by identifying infrastructure reliability as a critical adoption factor, a consideration often overlooked in implementations from high-resource settings. Our findings also highlight the necessity of guideline localization for clinical decision support systems. The misalignment between LLM recommendations and local protocols created some barriers to adoption and clinical utility, which should be resolved in future iterations of the product. Additionally, our results highlight important considerations regarding cost and resource constraints. Clinicians reported adjusting AI Consult recommendations based on the affordability and availability of medications for their predominantly low-income patient population.

This dimension of "contextual appropriateness" extends beyond technical accuracy to include socioeconomic feasibility, an important consideration rarely addressed in AI consult implementation literature from high-resource settings.

## Supporting information

Data collection tools and a results table

STROBE checklist compliance table

## Declarations

### Data Availability Statement

De-identified quantitative data are available from the corresponding author upon reasonable request. Raw qualitative data cannot be shared publicly due to data protection-related constraints (i.e., it includes disclosive information), but thematic analysis summaries are available upon request.

### Patient and Public Involvement Statement

Patients were not directly involved as research participants. Patient perspectives were incorporated through analysis of satisfaction metrics and feedback collected during routine care. All patient data were anonymized and analyzed in aggregate form.

### Author Contributions Statement

BAM secured the funding. CO conceptualized the study. BT and CO developed the methodology. GN, DO, EJ, and SW collected data. DO, GN, and SW performed data analysis. GN, DO, SW, EJ, and BAM drafted the original manuscript. All authors contributed to review and editing. BT, MEF, BAM, and CO supervised the project.

## Acknowledgments

We thank the leadership and staff of Penda Health for their collaboration and support. We especially acknowledge the 42 healthcare professionals who participated in this study. We also thank the technical team at Penda Health for providing access to system data and supporting data collection.

