## Supplementary material for "A Mixed-Methods Evaluation of Clinician Experiences and Adoption Patterns of an EHR-integrated Generative AI-based Clinical Decision Support in Kenya": Data collection tools and a results table

**Table of Contents**

1) Journey Mapping Interview Guide
2) User Experience Interview Guide
3) Focus Group Discussion Guide
4) Comprehensive Prompt
5) sTable 1: Total Number of Monthly Visits to Penda Health Facilities, and Proportion Where the ‘AI Consult’ Was Used

**Journey Mapping Interview Guide**

**Objective**: The purpose of this study is to explore how HCWs at Penda Health use AI at different touchpoints in their workflow, identify the factors that influence AI adoption, and uncover any challenges or barriers they encounter. The insights gathered will contribute to crafting the clinical data journey map and inform the development of interview and FGD guides.

**Background Questions**

- Can you describe where and how AI is used throughout the patient journey, and at which points you interact with it the most? Sketch the journey
- How does AI support your decision-making, and are there specific clinical decisions where it is particularly useful?
- What AI tools do you use, and how well do they integrate with your existing clinical workflow and systems?
- How does AI impact collaboration with other healthcare workers during patient care?
- How does AI assist in managing and interpreting patient information across different stages of care?
- What challenges or barriers do you face when using AI at various stages in the clinical journey?

**Touchpoint 1a: Pre-visit**

- Is there any customer interaction with Penda AI before interacting with Penda Health? Grace Health?

**Touchpoint 1b: Patient Triage (Initial Assessment)**

- Do you use AI tools during the initial patient triage or assessment process? If yes, how do you use them?
- Can you describe how AI supports or enhances your decision-making during patient triage?
- What are the key factors or conditions that make you decide to use AI tools during triage?
- What challenges do you face when using AI for patient triage?
- Are there limitations in the AI tools that affect their accuracy or usefulness during triage?

**Touchpoint 2: Waiting Areas**

- Do you use AI in the waiting areas? If so how?

**Touchpoint 3: Consultations**

- How do you use AI tools during patient consultations?
- In what ways does AI enhance the consultation process, such as in patient assessment or generating insights?
- Are there any specific AI influences your interaction with patients?
- What challenges or limitations do you encounter when using AI during consultations?

**Touchpoint 4: Diagnosis**

- Do you use AI to assist in diagnosing patients? If yes, how does the AI tool fit into your diagnostic process?
- Can you give an example of a case where AI influenced the diagnosis?
- What makes you decide to rely on AI for diagnosing a patient? Is it specific types of cases or symptoms?
- Do you trust AI-generated insights? Why or why not?
- What are the biggest challenges in using AI for diagnosis? (e.g., interpretability, accuracy, data quality, time)
- Have there been instances where AI suggested an incorrect diagnosis?

**Touchpoint 4: Treatment plan**

- Do you use AI tools to help in creating or personalizing treatment plans? If yes, how do you integrate AI insights into your decision-making process?
- Can you describe how AI contributes to optimizing treatment plans?
- In what types of cases or conditions do you find AI most helpful in treatment planning?
- Are there specific scenarios where you feel more comfortable using AI?
- What challenges or barriers do you encounter when using AI for treatment planning? (e.g., trust in recommendations, system complexity)
- Do you let patients know that AI was part of their treatment plan?

**Touchpoint 5: Pharmacy**

- Is AI utilized in pharmacy settings to support medication management and dispensing?
- In what ways does AI assist in verifying prescriptions or identifying potential drug interactions?
- Are there specific AI tools that help you in managing inventory or predicting medication demand? If so, how do they function?
- How does AI contribute to personalized medication plans or dosages based on patient data?
- What challenges do you encounter when using AI in pharmacy, such as concerns about accuracy, data privacy, or integration with existing systems?

**Touchpoint 6: Monitoring and Follow ups**

- Do you use the AI tool for monitoring patient progress after treatment or during follow-up visits? If yes, how do you use them?
- How does AI help in identifying patient outcomes or suggesting follow-up actions?
- What factors influence your decision to rely on AI for patient monitoring? Are there particular types of cases where AI is more useful?
- How does AI integrate with other monitoring systems or tools you use?
- What difficulties do you face when using AI to monitor patients? (e.g., technical issues, data accuracy, patient compliance)
- Are there concerns about the reliability of AI in tracking long-term health outcomes?

**User Experience Interview Guide**

**Introduction**

1. Welcome and Self-Introduction

- Greet the participant warmly and introduce yourself by name and role.
- Thank them for their time and acknowledge their participation.

2. Explain the purpose and key objectives of the interview

- Clarify that the goal is to understand their experience using the tool, including usability, workflow integration, accuracy, and areas for improvement.
- State the key objectives of the interview
  - Evaluate the usability of the AI consult tool.
  - Understand how it integrates into clinical workflows and affects efficiency.
  - Assess trust and accuracy in AI-generated recommendations.
  - Identify challenges and pain points faced by clinicians using the tool.
  - Gather suggestions for improvement from end-users

3. Encourage Honest and open Feedback

- Reassure the participant that there are no right or wrong answers.
- Emphasize that all feedback is valuable, whether positive or negative.
- Encourage them to provide specific examples where applicable

4. Set time expectations

- Let the participant know that the interview will take approximately 30–45 minutes.
- Reassure them that they are free to take a break or skip any questions if needed.

5. Obtain Consent

- Obtain verbal consent to record the session and written consent in case any photos will be taken.
- Confirm that participation is voluntary and that they can stop at any time.
- Address any questions from the participants before proceeding.

**Background Questions**

1. Clinician's Role and Experience

- Can you tell me about your role at Penda Health?
- How long have you been working as a clinician?
- How familiar are you with using AI tools in your clinical practice?

2. Use of the AI Consult Tool.

- How long have you been using the AI consult tool at Penda Health?
- How frequently do you use the AI consult tool in your daily workflow (e.g., every consultation, occasionally)?

**Usability and User Experience**

1. Initial Impressions

- What were your first impressions of the AI consult tool when you started using it?
- Did the tool meet your expectations when you first used it? Why or why not?

2. Ease of Use

- How easy or difficult is it to use the AI consult tool?
- Can you describe any specific features that you found particularly helpful or challenging?
- How quickly can you navigate through the tool during a consultation?
- Are there any points in the workflow where you feel the tool slows you down?
- tool?

3. Use of the Penda AI consult to generate different reports

a) Summary Reports

- Can you walk me through your experience using the AI tool to generate summary reports?
- Does the tool give you control over the level of detail in the summary? If so, how effective is it in tailoring the summary to your needs?
- How easy is it for you to review and edit AI-generated summaries before finalizing them? Are there any challenges in making adjustments?
- Based on your experience, how accurate and relevant are the AI-generated summaries? Do they capture the key points effectively?

b) For Comprehensive Reports:

- When generating comprehensive reports using the tool, how easy is it?
- Does the tool allow for customization in the report format or structure? If so, how useful is this feature for your reporting needs?
- How do you verify the accuracy of the data and insights presented in the AI-generated comprehensive reports? Does the tool assist in this process?
- Have you encountered any gaps or missing data when generating comprehensive reports? How does the tool help you address these issues?

c) For Treatment Plans (Healthcare context):

- When using the AI tool to generate treatment plans, how user-friendly is the interface for inputting patient data and receiving recommendations?
- Are you able to adjust the treatment plans based on your clinical judgment? If so, how well does the tool allow for these adjustments?
- How satisfied are you with the speed and accuracy of the AI-generated treatment plans? Can you provide specific examples?
- Does the AI tool explain how it arrived at its treatment recommendations? If so, do you find this level of transparency helpful?

d) On Ethics and Bias:

- How does the tool communicate potential biases or limitations in its treatment recommendations? Do you feel it handles these issues effectively?
- Is there an option to override or question the AI’s recommendations? How often do you find yourself using this feature?
- Does the tool offer any insights into the diversity of the data it uses to generate treatment plans, particularly for ensuring equitable outcomes across different patient groups?

4. Integration with Workflows and Human Expertise

- How does the AI consult tool fit into your existing workflow? Does it complement your clinical tasks or create disruptions?
- Are there any moments when you feel the tool integrates seamlessly with patient care? Can you provide an example?
- Have you encountered any difficulties or challenges when switching between the AI consult tool and other tools or systems you use during consultations?
- How well does the AI-generated treatment plan integrate into your workflow and electronic health records (EHR) system?
- Do you find it easy to cross-reference AI recommendations with your own expertise or clinical guidelines? How often do you rely on your judgment over the AI’s suggestions?
- Overall, how would you describe the balance between AI suggestions and human oversight when developing treatment plans? What improvements, if any, would you suggest?

5. Accuracy and Trust in AI Recommendations

- How accurate do you find the AI-generated recommendations or diagnostic support?
- Can you recall a situation where the AI consult tool's suggestions were particularly helpful or inaccurate?
- How do you respond when the AI tool provides a recommendation that seems incorrect or incomplete
- How much do you rely on the tool’s recommendations when making clinical decisions?
- Do you trust the AI’s output, or do you feel the need to double-check frequently?

**Interaction and User Interface**

1. Interface Design

- How would you rate the tool’s responsiveness? Does it load and respond quickly enough during consultations?

2. Help and Support

- Have you ever needed help or guidance while using the tool? If so, did you find the help options sufficient?
- Is there anything you think could be improved in terms of support or guidance within the tool?

**Impact on Patient Care and Outcomes**

1. Impact on Consultation Time

- How has the AI consult tool affected the duration of your patient consultations?
- Do you feel that the tool helps you save time, or does it add extra steps to the consultation process?

2. Impact on Patient Outcomes

- Do you believe the AI consult tool has positively impacted patient outcomes (e.g., more accurate diagnoses, better treatment plans)?
- Have patients responded positively or negatively to the use of AI during their consultation?

**Challenges and Improvements**

1. Pain Points

- What are the biggest challenges or frustrations you have encountered while using the AI consult tool?
- Have you experienced any technical issues, such as bugs or system crashes?

2. Suggestions for Improvement

- What specific improvements would you suggest to make the AI consult tool more user-friendly?
- Are there any features you would like to see added to the tool?
- If you could change one thing about the AI consult tool, what would it be?

**Final Thoughts**

1. Overall Experience

- How would you summarize your overall experience using the AI consult tool?
- On a scale from 1 to 10, how would you rate the tool’s usefulness in your clinical practice?

2. Closing Remarks

- Is there anything else you would like to add or any feedback you haven’t yet shared?
- Thank the clinician for their time and valuable feedback.

**Focus Group Discussion Guide**

**Objective:** To understand how facility managers monitor AI usage, its integration into workflows, and its impact on clinical outcomes.

**Section 1: Introduction**

1. Welcome and Objectives:
   - Thank participants for attending.
   - Explain the purpose: To explore how facility managers monitor AI usage, identify challenges, and recommend improvements for effective utilization at the facility level.
   - Emphasize confidentiality and the value of their input.
2. Participant Introductions:
   - Name, role, and experience with AI implementation and management in their facility.

**Section 2: Current AI Usage Practices**

1. AI Integration and Utilization:
   - What are the main tasks or areas where AI is used in your facility? (e.g., triage, consultation, decision-making, referrals)
   - How has AI been integrated into the existing EMR or other systems?
2. Training and Onboarding:
   - What training or support do staff receive to use AI effectively?
   - Are there refresher sessions or continuous training on AI updates and new functionalities?

**Section 3: Monitoring AI Usage**

1. Usage Tracking:
   - What mechanisms are in place to monitor AI usage among clinical staff? (e.g., reports, dashboards, audits)
   - Are you able to track how frequently AI is consulted and the types of cases it supports?
2. Performance Review:
   - How do you assess whether AI is being used correctly or effectively by staff?
   - Are there instances where AI usage is monitored for clinical outcomes or patient safety concerns?
3. Compliance and Reporting:
   - Do you review whether staff consistently use AI in cases where it is recommended? If so, how?
   - How do you address cases where AI is not used or its recommendations are ignored?
4. Feedback Collection:
   - Is there a formal process for staff to share feedback about AI's performance or utility?
   - How is this feedback incorporated into monitoring or improvement processes?

**Section 4: Challenges in Monitoring AI Usage**

1. System-Level Challenges:
   - What challenges do you face in monitoring AI usage at the facility level? (e.g., technical issues, lack of reporting tools, resistance from staff)
   - Are there any gaps in the data generated by AI or the reports available to managers?
2. Staff-Level Challenges:
   - How do you ensure staff adhere to AI recommendations while maintaining clinical judgment?
   - Are there challenges with staff perception of AI (e.g., trust, over-reliance, or resistance)?
3. Resource Constraints:
   - Are there resource limitations (e.g., IT support, training time) that impact AI monitoring efforts?

**Section 5: Recommendations for Effective Monitoring**

1. Improving Monitoring Processes:
   - What tools or processes would help you better monitor AI usage at the facility level?
   - How could dashboards, reports, or audits be enhanced to provide actionable insights?
2. Enhancing Staff Engagement:
   - What strategies could improve staff compliance with AI usage while respecting their clinical expertise?
   - How can facility managers support staff in resolving challenges with AI integration or usage?
3. Strengthening Feedback Loops:
   - How can feedback from staff about AI be better collected, analyzed, and used to improve its functionality?
   - What role should facility managers play in communicating staff concerns or suggestions to developers?

**Section 6: Closing**

1. Summary of Key Points:
   - Recap major insights and suggestions shared during the discussion.
2. Open Floor for Final Thoughts:
   - Is there anything else you’d like to share about monitoring AI usage at the facility level?
3. Thank You and Next Steps:
   - Thank participants for their valuable contributions.
   - Share how their insights will guide improvements in AI monitoring practices.

**Comprehensive Prompt**

“You are a consultant physician who acts as a supportive mentor to clinical officers in an urgent care setting in Nairobi. The clinical officer will provide you with details about the patient's case they are seeing and you should provide easy-to-read feedback. The type of feedback you give must be context-dependent. For example:

- If the condition is low-acuity and you don't see any concerning vital signs or clinical findings, such as a patient with common cold symptoms and normal vital signs, you can provide brief positive feedback on documentation and perhaps an interesting clinical teaching point relevant to the case and brief suggestions for symptomatic treatment such as throat lozenges.
- In contrast, if the documentation shows that the clinician could be missing something more serious, such as a patient with severe headache, fever, and stiff neck, the AI feedback should be more directive, such as, "This patient potentially has meningitis" and provide diagnostic and treatment guidance accordingly.
- At times, the documentation may show errors, for example, a child with viral gastroenteritis who is prescribed antibiotics. The feedback should be more directive saying: “hold on, there appears to be a mismatch between your treatment plan and the diagnosis given. Viral gastroenteritis requires a careful assessment of hydration status and the provision of zinc and ORS”.

In the end, your brief feedback should be highly practical, relevant, and easy for clinicians to read and act on. The tone should be more conversational and personal. Please be concise in your response.”

**sTable 1: Total Number of Monthly Visits to Penda Health Facilities, And Proportion Where the ‘AI Consult’ Was Used**

| Months | Total Number of Visits | Number of Visits where ‘AI Consult’ Was Used | % ‘AI Consult’ Usage | % MoM AI Usage Change |
| --- | --- | --- | --- | --- |
| Feb | 31,405 | 1,391 | 4% | - |
| Mar | 34,961 | 2,662 | 8% | 91.37 |
| Apr | 29,823 | 2,428 | 8% | -8.79 |
| May | 32,340 | 4,082 | 13% | 68.12 |
| Jun | 33,852 | 6,880 | 20% | 68.54 |
| Jul | 33,926 | 11,215 | 33% | 63.01 |
| Aug | 30,050 | 12,549 | 42% | 11.89 |
| Sep | 31,749 | 14,843 | 47% | 18.28 |
| **Totals** | **258,106** | **56,050** |  |  |

Legend: MoM = Month on Month
