## Supplementary material for "A Mixed-Methods Evaluation of Clinician Experiences and Adoption Patterns of an EHR-integrated Generative AI-based Clinical Decision Support in Kenya": STROBE checklist compliance table

Appendix: STROBE Checklist Compliance Table

**STROBE Checklist for Observational Study Components**

| Section /Item | Item No. | Recommendation | Addressed in Manuscript | Notes for Team |
| --- | --- | --- | --- | --- |
| Title and abstract | 1 | (a) Indicate the study's design with a commonly used term in the title or the abstract | Yes - Abstract clearly identifies this as a "mixed-methods study" | Consider adding "observational" to clarify design |
|  |  | (b) Provide in the abstract an informative and balanced summary of what was done and what was found | Yes - Abstract includes background, objective, methods, results and conclusions |  |
| Introduction |  |  |  |  |
| Background /rationale | 2 | Explain the scientific background and rationale for the investigation being reported | Yes - Introduction section 1.1 |  |
| Objectives | 3 | State specific objectives, including any prespecified hypotheses | Yes - Section 1.3 | Were there any specific hypotheses that should be stated? |
| Methods |  |  |  |  |
| Study design | 4 | Present key elements of study design early in the paper | Yes - Section 2.1 |  |
| Setting | 5 | Describe the setting, locations, and relevant dates, including periods of recruitment, exposure, follow-up, and data collection | Yes - Section 2.1 and 2.4 |  |
| Participants | 6 | (a) Give the eligibility criteria, and the sources and methods of selection of participants | Partial - Section 2.3 | Need more detail on specific inclusion/exclusion criteria for participants |
| Variables | 7 | Clearly define all outcomes, exposures, predictors, potential confounders, and effect modifiers | Partial - Section 2.5 | Need more explicit definition of variables used in quantitative analysis |
| Data sources /measurement | 8 | For each variable of interest, give sources of data and details of methods of assessment | Yes - Section 2.4 |  |
| Bias | 9 | Describe any efforts to address potential sources of bias | Partial - Section 2.3 mentions random sampling to minimize selection bias | What other bias mitigation strategies were used? |
| Study size | 10 | Explain how the study size was arrived at | Partial - Section 2.3 describes selection of facilities and participants | Was a power calculation or other formal determination of sample size conducted for quantitative components? |
| Quantitative variables | 11 | Explain how quantitative variables were handled in the analyses | Partial - Section 2.5 | More detail needed on handling of quantitative variables |
| Statistical methods | 12 | (a) Describe all statistical methods, including those used to control for confounding | Yes - Section 2.5 |  |
|  |  | (b) Describe any methods used to examine subgroups and interactions | Partial - Section 2.5 | Additional detail would strengthen this section |
|  |  | (c) Explain how missing data were addressed | No | Does the manuscript address how missing data were handled? |
|  |  | (d) If applicable, explain how loss to follow-up was addressed | Not applicable | No follow-up component in this cross-sectional study |
|  |  | (e) Describe any sensitivity analyses | Partial - HLM mentioned in results | Consider adding details on sensitivity analyses if conducted |
| Results |  |  |  |  |
| Participants | 13 | (a) Report numbers of individuals at each stage of study | Yes - Section 3 introduction |  |
|  |  | (b) Give reasons for non-participation at each stage | No | Were there any declined invitations or dropouts? |
|  |  | (c) Consider use of a flow diagram | No | A participant flow diagram would strengthen the manuscript |
| Descriptive data | 14 | (a) Give characteristics of study participants | Yes - Table 1 and Section 3 |  |
|  |  | (b) Indicate number of participants with missing data for each variable of interest | No | Was there missing data? |
| Outcome data | 15 | Report numbers of outcome events or summary measures | Yes - Sections 3.1-3.4 |  |
| Main results | 16 | (a) Give unadjusted estimates and, if applicable, confounder-adjusted estimates and their precision | Yes - Sections 3.1.3, 3.2, and 3.4 |  |
|  |  | (b) Report category boundaries when continuous variables were categorized | Partial | Some explanation of categorization in user types, but could be clearer |
|  |  | (c) If relevant, consider translating estimates of relative risk into absolute risk | Not applicable |  |
| Other analyses | 17 | Report other analyses done—e.g. analyses of subgroups and interactions, and sensitivity analyses | Yes - Sections 3.1.3 and 3.2 |  |
| Discussion |  |  |  |  |
| Key results | 18 | Summarise key results with reference to study objectives | Yes - Section 4.1 |  |
| Limitations | 19 | Discuss limitations of the study, taking into account sources of potential bias or imprecision | Yes - Section 4.5 |  |
| Interpretation | 20 | Give a cautious overall interpretation of results considering objectives, limitations, multiplicity of analyses, results from similar studies, and other relevant evidence | Yes - Sections 4.2, 4.3, 4.4 |  |
| Generalisability | 21 | Discuss the generalisability (external validity) of the study results | Yes - Section 4.5 and 4.6 |  |
| Other information |  |  |  |  |
| Funding | 22 | Give the source of funding and the role of the funders for the present study | Yes - Funding statement in Declarations |  |

Appendix: SRQR Checklist Compliance Table

| Section | Item | Description | Addressed in Manuscript | Notes for Team |
| --- | --- | --- | --- | --- |
| Title and abstract |  |  |  |  |
| Title | S1 | Concise description of the nature and topic of the study | Yes |  |
| Abstract | S2 | Summary of key elements of the study using the abstract format of the intended publication | Yes |  |
| Introduction |  |  |  |  |
| Problem formulation | S3 | Description and significance of the problem/phenomenon studied | Yes - Section 1.1 |  |
| Purpose or research question | S4 | Purpose of the study and specific objectives or questions | Yes - Section 1.3 |  |
| Methods |  |  |  |  |
| Qualitative approach and research paradigm | S5 | Qualitative approach and theory, philosophical or conceptual framework | Yes - Section 2.2 |  |
| Researcher characteristics and reflexivity | S6 | Researcher characteristics that may influence the research | No | How was researcher positionality addressed? |
| Context | S7 | Setting/site and contextual factors | Yes - Section 2.1 |  |
| Sampling strategy | S8 | How and why research participants were selected and sample size | Partial - Section 2.3 | More detail on qualitative sampling rationale needed |
| Ethical issues | S9 | IRB/ethics committee approval, ethical issues in the study | Yes - Section 2.6 and Declarations |  |
| Data collection methods | S10 | Types of data collected, protocol, instruments used | Yes - Section 2.4 |  |
| Data collection instruments and technologies | S11 | Instruments and devices used for data collection | Yes - Section 2.4 |  |
| Units of study | S12 | Number and relevant characteristics of participants, documents, or events included in the study | Yes - Section 3 |  |
| Data processing | S13 | Methods for processing data prior to and during analysis | Yes - Section 2.5 |  |
| Data analysis | S14 | Process of examining data and producing findings | Yes - Section 2.5 |  |
| Techniques to enhance trustworthiness | S15 | Techniques to enhance trustworthiness and credibility of data analysis | Partial - Section 2.5 mentions member checking | What other trustworthiness strategies were employed? |
| Results/findings |  |  |  |  |
| Synthesis and interpretation | S16 | Main findings and integration of findings | Yes - Sections 3.2, 3.3, 3.4 |  |
| Links to empirical data | S17 | Evidence (quotes, field notes, text excerpts, photographs) to substantiate analytic findings | Yes - Quotations included throughout Section 3 |  |
| Discussion |  |  |  |  |
| Integration with prior work, implications, transferability, and contribution to the field | S18 | How findings fit with existing literature, relevance to policy and practice | Yes - Section 4.2 |  |
| Limitations | S19 | Limitations of the study | Yes - Section 4.5 |  |
| Other |  |  |  |  |
| Conflicts of interest | S20 | Potential sources of influence on study conduct and conclusions | Yes - Competing interests statement |  |
| Funding | S21 | Sources of funding and role of funders in the study | Yes - Funding statement |  |
